# Mitigating the psychological impacts of COVID-19 restrictions: The <u>B</u>ehavioural <u>A</u>ctivation in <u>S</u>ocial <u>I</u>so<u>l</u>ation (BASIL) pilot randomised controlled trial to prevent depression and loneliness among older people with long term conditions

**DOI:** 10.1101/2021.05.17.21257309

**Authors:** Simon Gilbody, Elizabeth Littlewood, Dean McMillan, Carolyn A. Chew-Graham, Della Bailey, Samantha Gascoyne, Claire Sloan, Lauren Burke, Peter Coventry, Suzanne Crosland, Caroline Fairhurst, Andrew Henry, Catherine Hewitt, Kalpita Joshi, Eloise Ryde, Leanne Shearsmith, Gemma Traviss-Turner, Rebecca Woodhouse, Andrew Clegg, Tom Gentry, Andrew Hill, Karina Lovell, Sarah Dexter Smith, Judith Webster, David Ekers

**Affiliations:** Department of Health Sciences, University of York, York, YO10 5DD, UK; Hull York Medical School, University of York, York, YO10 5DD, UK; School of Medicine, Keele University, Staffordshire, ST5 5BG, UK; Tees, Esk and Wear Valleys NHS FT, Research & Development, Flatts Lane Centre, Middlesbrough, TS6 0SZ, UK; Leeds Institute of Health Sciences, University of Leeds, Leeds, LS2 9NL, UK; Age UK, Tavis House, 1-6 Tavistock Square, London WC1H 9NA; Division of Nursing, Midwifery & Social Work, University of Manchester, Oxford Road, Manchester, M13 9PL; Patient and Public Representative, UK

## Abstract

**Background:** Older adults with long-term conditions have become more socially isolated (often due to advice to ‘shield’ to protect them from COVID-19) and are thus at particular risk of depression and loneliness. There is a need for brief scalable psychosocial interventions to mitigate the psychological impacts of social isolation. Behavioural Activation is a plausible intervention, but a trial is needed.

**Methods:** We undertook an external randomised pilot trial (ISRCTN94091479) designed to test recruitment, retention and engagement with, and the acceptability and preliminary effects of the intervention. Participants aged ≥ 65 years with two or more long-term conditions were recruited between June and October 2020. Behavioural Activation was offered to intervention participants (n=47), and control participants received usual care (n=49).

**Findings:** Remote recruitment was possible and 45/47 (95.7%) randomised to the intervention completed one or more sessions (median 6 sessions). 90 (93.8%) completed the one month follow-up, and 86 (89.6%) completed the three month follow-up. The between-group comparison for the primary clinical outcome at one month was an adjusted between group mean difference of −0.50 PHQ-9 points (95% CI −2.01 to 1.01), but only a small number of participants had completed the intervention at this point. At three months, the PHQ-9 adjusted mean difference was 0.19 (95% CI −1.36 to 1.75). When we examined loneliness, the between-group difference in the De Jong Gierveld Loneliness scale at one month was 0.28 (95% CI −0.51 to 1.06), and there was statistically significant between group difference at three months (−0.87; 95% CI −1.56 to −0.18). Participants who withdrew had minimal depressive symptoms at entry.

**Interpretation:** Behavioural Activation is a plausible intervention to mitigate the psychological impacts of COVID-19 isolation for older adults. The intervention can be delivered remotely and at scale, but should be reserved for older adults with evidence of depressive symptoms. The significant reduction in loneliness is unlikely to be a chance finding, and this will now be confirmed in a fully powered RCT.

**Funding:** This study was funded by National Institute for Health Research (NIHR) Programme Grants for Applied Research (PGfAR) RP-PG-0217-20006

## Introduction

In March 2020 a pandemic due to a new virus, the Acute Respiratory Syndrome Coronavirus 2 (SARS-CoV-2), was declared. The first wave reached the United Kingdom (UK) within a short period of time and in March 2020 the UK governments (including devolved nations) administered a national Stay At Home order (“lockdown”), which included instructions for people to follow social distancing and self-isolation guidelines, and recommendations for strict isolation (“shielding”) for the most vulnerable (such as those with long-term conditions and older people) in order to protect their own and others’ health, and to avoid a sudden increase in demand on the NHS. Shielding orders were eased in the second half of 2020 but were reintroduced in January 2021 as part of a further lockdown in response to subsequent COVID-19 waves. Many people with long-term conditions have remained avoidant of social contact in order to protect themselves from COVID-19 throughout the pandemic, irrespective of official guidance.^1^ Similar recommendations and restrictions were also set in place in many health care systems around the world.

The mental health of the population has deteriorated during COVID-19.^2^ Many report social isolation, and the incidence of depression and anxiety have increased for older people and those with medical vulnerabilities.^3^ A plausible mechanism for this is that COVID-19 restrictions have led to disruption of daily routine, loss of social contact and heightened isolation and increased loneliness, which are each powerful precipitants of mental ill health.^4^ Anticipating these behavioural and psychological consequences, a rapid review published in The Lancet^5^ highlighted the detrimental impacts on mental health of quarantine, but offered limited advice on how this could be mitigated.

Social isolation, social disconnectedness, perceived isolation and loneliness are known to be linked to common mental health problems, such as depression in older people.^4^ The impairments in quality of life associated with depression are comparable to those of major physical illness.^6^ Loneliness is a risk factor for depression and is also known to be detrimental to physical health and life expectancy.^7,8^

Loneliness is not an inevitable consequence of social isolation and strategies to prevent or mitigate loneliness were recognised as a population priority even before COVID-19.^9,10^ There are a number of promising interventions that focus on using social networks^11^ or adapting the strategies central to cognitive behavioural therapy.^12^ It is recognised that strategies that, for instance, maintain social connectedness could be important in ensuring the population mental health of older people,^13^ particularly during the pandemic^4^ and in the planning for post-pandemic recovery.^14^ If a brief intervention for depression and loneliness could be delivered at distance (such as via telephone) and at scale, then this would lead to significant benefits to the NHS and society. This could potentially mitigate the immediate and longer lasting psychological impacts of COVID-19 on vulnerable populations, including older people and those with long-term conditions.^15^

Our research collaborative has previously developed, with input from older adults and carers, a credible intervention that can potentially meet these needs in populations of older people,^16^ and we have evaluated this in older populations with high rates of multiple long term conditions.^17,18^ Behavioural Activation (BA) is a practical treatment that explores how physical inactivity and low mood are linked, and result in a reduction of valued activity.^19^ Within BA, the therapist and patient work together to develop a collaborative treatment plan that seeks to reinstate (or replace, if former activities are no longer possible) behaviours that connect people to sources of positive reinforcement (meaningful activity), including social connectedness. However, this has not yet been tested in a large-scale clinical trial, or in the context of the COVID-19 pandemic where social isolation is more prevalent. Small scale trials of BA delivered to socially-isolated older people have produced encouraging preliminary results,^20^ but there is not yet sufficient research evidence to support whole-scale adoption, or to inform the population response to COVID-19.

Along with many researchers working in the field of mental health, we were keen to use our existing research expertise and research capacity to help mitigate the impact of the COVID-19 pandemic. We therefore adapted our existing NIHR-funded programme of work in early-2020 to answer the following question: **‘Can we prevent or ameliorate depression and loneliness in older people with long term conditions during isolation?’**.

In this paper we present the rationale and results of a pilot randomised controlled trial of manualised BA, adapted specifically to be delivered at scale and remotely (via the telephone or video call) for older adults who may have become socially isolated as a consequence of COVID-19.

## Methods

### Study design and participants

BASIL is an external pilot randomised controlled trial (RCT)^21^ and includes a concurrent qualitative study. The BASIL pilot is designed to provide key information on methods of recruitment, intervention uptake, retention, experience of the BA intervention for our target population, and acceptability of the intervention and training for intervention practitioners (hereafter BASIL Support Workers). Here we report the preliminary results and key adaptations for use in COVID-19 and older people with long term conditions, alongside the preliminary results of comparative effectiveness.

The COVID-19 responsive BASIL trials programme is supported by the National Institute for Health Research (NIHR) under grant RP-PG-0217-20006, and was adopted by the NIHR Urgent Public Health programme on 28^th^ May 2020 (https://www.nihr.ac.uk/covid-studies/study-detail.htm?entryId=249030). The protocol for the BASIL pilot study was pre-registered (ISRCTN94091479) on 9^th^ June 2020 and recruitment took place between 23^rd^ June and 15th October 2020 (18 weeks in total). Older adults at risk of loneliness and depression as a consequence of social isolation under COVID-19 restrictions were recruited from primary care registers. They were randomised to receive either usual primary care from their general practice or Behavioural Activation intervention in addition to usual care (see below for full description of usual care and BA intervention).

#### Inclusion criteria

Older adults (65 years or over) with two or more physical long-term conditions (LTCs). The pragmatic definition and type of LTCs mirror that applied in primary care in the UK ^22^ and we focussed on common LTCs experienced by older people (such as asthma/COPD, diabetes, hypertension/coronary heart disease, stroke) according to the primary care Quality and Outcomes Framework (QOF),^23^ but also included conditions such as musculoskeletal problems and chronic pain. Participants included those subject to Government guidelines regarding COVID-19 self-isolation, social distancing and shielding as relevant to their health conditions and age (though this was not a requirement and these requirements changed during the study period).

#### Exclusion criteria

Older adults who have cognitive impairment, bipolar disorder/psychosis/psychotic symptoms, alcohol or drug dependence, in the palliative phase of illness, have active suicidal ideation, are currently receiving psychological therapy, or are unable to speak or understand English.

Potentially eligible patients were contacted by telephone by staff working with the general practices. Those patients who expressed an interest in the study during this initial telephone contact provided their verbal ‘permission to contact’ for a member of the study team to contact them by telephone to discuss the study and determine eligibility. Interested patients could also complete an online consent form or contact the study team directly.

### Randomisation, concealment of allocation and masking

After consent, eligible participants completed a baseline questionnaire over the telephone with a study researcher. Participants were then randomised and informed of their group allocation (intervention or usual care with signposting). Participants were allocated in a 1:1 ratio using simple randomisation without stratification. Treatment allocation was concealed from study researchers at the point of recruitment using an automated computer data entry system, administered remotely by the York Trials Unit and using a computer-generated code. Owing to the nature of the intervention, none of the participants, general practices, study clinicians, or BASIL Support Workers could be blinded to treatment allocation. GPs were informed by letter of participant treatment allocation. Outcome assessment was by self-report, and study researchers facilitating the telephone-based outcome assessment were blind to treatment allocation.

### Intervention (Behavioural Activation)

The intervention (Behavioural Activation within a collaborative care framework) has been described elsewhere^17^ and was adapted for the purposes of the BASIL trial. Within the BASIL Behavioural Activation intervention, the therapist (‘BASIL Support Worker’ (BSW)) and participant worked together to develop a collaborative treatment plan that sought to reinstate (or replace, if former activities were no longer possible because of social isolation and/or long-term conditions) behaviours that connect them to sources of positive reinforcement (valued activity). BA has the potential to address depression and loneliness in the presence of social-isolation in this way^16,24^ and the simplicity of BA made it suitable for delivery in the context of COVID-19.

Intervention participants were offered up to eight sessions over a 4 to 6 week period delivered by trained BSWs, accompanied by participant materials. Participants in the intervention group were provided with a BASIL Behavioural Activation workbook. This booklet was modified to take account of Government guidance regarding the need for social isolation/physical distancing and enforced isolation for those people most at risk (‘extremely vulnerable’ people). For example, the BASIL booklet discussed ways to replace activities which are no longer possible with ones which preserve social distancing whilst helping participants stay connected with the activities and people important to them; illustrative patient stories included in the booklet were modified to take account of COVID-19 restrictions. Behavioural Activation acknowledged the disruption to people’s lives and usual routines and encouraged the establishment of a balanced daily routine. The intervention also recognised that participants may be worried about the current situation due to COVID-19 and suggested strategies to help cope.

All intervention sessions were delivered remotely via telephone or video call, according to participant preference. The first session was scheduled to last approximately one hour, with subsequent sessions lasting approximately 30 minutes.

Depression symptom monitoring at each intervention session was undertaken using a validated depression scale (the DASS^25^) with scores guiding decision-making by BSWs, and guided by supervision provided by clinical members of the study team. Where risk or significant clinical deterioration was noted the participant was supported to access more formal healthcare interventions. Where feasible and where considered appropriate and acceptable by the participant and BSW, the intervention was extended to include involvement of a participant’s informal caregiver/significant other. Intervention participants continued to receive their usual care/treatment (where this was feasible given COVID-19) alongside the BASIL intervention and no treatment was withheld..

### Comparator (usual GP care)

Participants in the control group received usual care as provided by their current NHS and/or third sector providers. In addition, control participants were ‘signposted’ to reputable sources of self-help and information, including advice on how to keep mentally and physically well. Examples of such sources was the Public Health England (PHE) ‘Guidance for the public on the mental health and wellbeing aspects of coronavirus (COVID-19)’^26^ and Age UK.^27^

### Outcome measures

Demographic information was obtained at baseline and included: age, gender, LTC type, socio-economic status, ethnicity, education, marital status, and number of children.

Outcome measures were collected at baseline, one, three and 12 months post-randomisation. The PHQ-9 was also applied at screening to ascertain risk of self-harm or suicide. The primary clinical outcome was self-reported symptoms of depression, assessed by the PHQ-9,^28^ and the primary time point was one month. We also measured PHQ-9 depression severity at three and 12 months post-randomisation. Other secondary outcomes measured at one, three and 12 months were health related quality of life (measured by the SF-12v2 mental component scale (MCS) and physical component scale (PCS)),^29^ anxiety (measured by the GAD-7),^30^ perceived social and emotional loneliness (measured by the De Jong Gierveld Scale −11 items loneliness scale) and questions relating to COVID-19 circumstances and adherence to government guidelines.^31^ We only report outcomes at one and three months since these timepoints informed the full trial design, and the BASIL trial participants remain in follow up for their most distal outcome.

### Sample size & statistical analysis

#### Sample size

The primary aim of the BASIL pilot trial was to test the feasibility of the intervention and the methods of recruitment, randomisation and follow-up.^21^ Sample size calculations were based on estimating attrition and standard deviation (SD) of the primary outcome. We aimed to recruit 100 participants. The intervention was delivered by BSWs and allowed for potential clustering by BSWs assuming an inter-cluster correlation (ICC) of 0.01 and average cluster size of 15 based upon previous studies.^17^ The effective sample size was therefore 88. Anticipating 15-20% of participants would be lost to follow-up (17% in the CASPER trial of older adults^17^), this would result in an effective sample size of at least 70 participants which is sufficient to allow reasonably robust estimates of the SD of the primary outcome measure to inform the sample size calculation for a definitive trial.^32,33^

#### Statistical analysis

The flow of participants through the pilot trial (number of people identified, approached, screened, eligible, randomised, receiving the intervention, and providing outcome data) is detailed in a CONSORT flow diagram as per pilot trial recommendations.^21^ The number of individuals withdrawing from the intervention and/or the trial, and any reasons for withdrawal, was summarised by trial arm. All baseline and outcome data were summarised descriptively, by trial arm, using mean and standard deviation (SD) for continuous outcomes, and count and percentage for categorical data. To quantify the acceptability of the intervention the number and duration of sessions were summarised.

In our prespecified statistical analysis plan, linear regression was used to explore differences in the PHQ-9 and De Jong Gierveld Loneliness Scale, adjusting for the baseline measure of the score, between groups at one and three months. The adjusted mean difference and 95% confidence interval (CI) was reported as preliminary estimates of effect but this pilot trial was not powered to show efficacy.

### Process evaluation

A nested qualitative study was conducted to provide important learning about the study processes, and acceptability of the BASIL intervention. We planned semi-structured interviews of up to 15 participants who completed the BASIL intervention (‘completers’), up to 10 participants who did not complete the intervention (‘non-completers’) and all BSWs who delivered the intervention (n=9). Interviews explored views and experiences of the study and acceptability of the intervention. Initial thematic analysis^34^ and subsequent analysis sensitised by the Theoretical Framework of Acceptability ^35^ was undertaken.

### Patient and Public Involvement (PPI)

The BASIL trial was informed by a Patient and Public Involvement Advisory Group (PPI AG) who were working with the research collective on the existing NIHR-funded research programme. This PPI AG included older adults with lived experience of mental health and/or physical health conditions, and caregivers. The PPI AG were consulted on many aspects of the trial design including modification of the BA intervention for BASIL, remote recruitment of BASIL participants, and the relevance and readability of study recruitment information. The group are a vital component of the BASIL trials programme and will continue to contribute throughout the delivery of this work.

## Results

### Participant recruitment and characteristics

Two general practices within Tees, Esk and Wear Valleys NHS Foundation Trust conducted database searches and mailouts. 799 study information packs were mailed out across the two practices between 17th June and 4th September 2020 (initially in batches of 50, later increased to 100). Of these 104 were screened for eligibility and 96 were recruited out of our target of 100: three were not eligible as they were currently in receipt of psychological treatment, two eligible participants did not complete the baseline questionnaire following consent, and three others were not randomised for other reasons. Participants were randomised between 23rd June and 15th October 2020: 47 to the BA intervention group; and 49 to usual care with signposting group (Figure 1).

**Figure 1:**
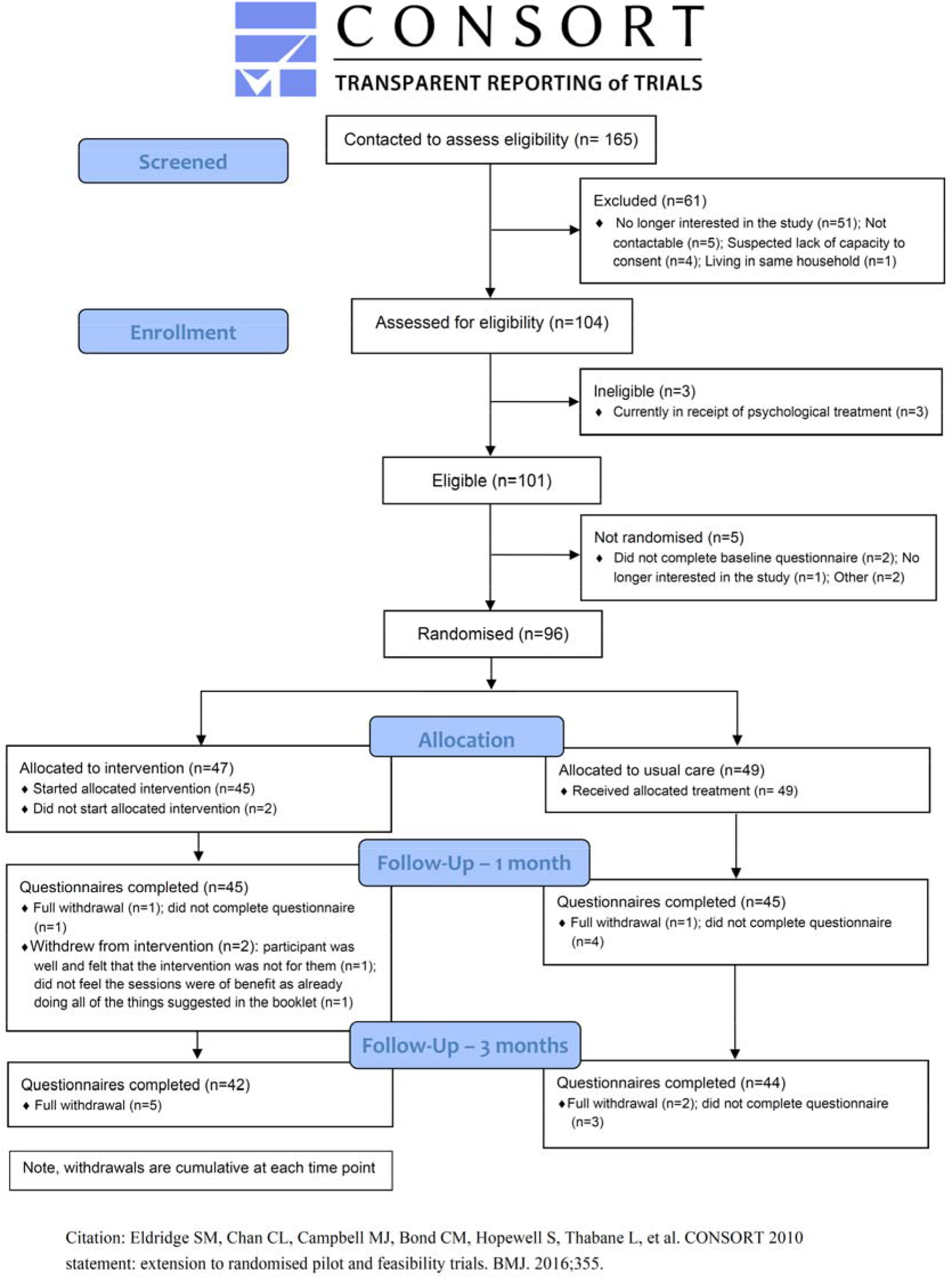
BASIL CONSORT flow diagram.

On average, participants were aged 74 years (SD 5.5), and were mostly White (n=92, 95.8%) and approximately two-thirds of the sample were female (n=59, 61.5%) (Table 1). Cardiovascular conditions (49.0%) and arthritis (38.5%) were the most commonly reported long term health conditions. The majority of participants (55.2%) were social/physical distancing and adhering to UK Government’s guidance in relation to COVID-19 restrictions all of the time (65.6%). There was reasonable balance in baseline characteristics between the two groups, but with some differences including a larger proportion of females, and current and former smokers, and fewer participants shielding in the usual care group than the intervention group.

**Table 1:** Baseline demographics of participants as randomised

| Demographic |  | Usual<br>Care<br>(n=49) | Intervention<br>(n=47) | Total<br>(n=96) |
| --- | --- | --- | --- | --- |
| Age, years | Mean (SD) | 74.1 (5.6) | 74.2 (5.4) | 74.2 (5.5) |
| Gender, n (%) | Male | 17 (34.7) | 20 (42.6) | 37 (38.5) |
|  | Female | 32 (65.3) | 27 (57.4) | 59 (61.5) |
| Ethnicity, n (%) | White | 47 (95.9) | 45 (95.7) | 92 (95.8) |
|  | Black or Black British | 0 (0.0) | 0 (0.0) | 0 (0.0) |
|  | Asian or Asian British | 0 (0.0) | 0 (0.0) | 0 (0.0) |
|  | Other | 2 (4.1) | 2 (4.3) | 4 (4.2) |
| LTC type, n (%) | Cardiovascular condition | 21 (42.9) | 26 (55.3) | 47 (49.0) |
|  | Arthritis | 21 (42.9) | 16 (34.0) | 37 (38.5) |
|  | Respiratory condition | 17 (34.7) | 18 (38.3) | 35 (36.5) |
|  | Diabetes | 14 (28.6) | 14 (29.8) | 28 (29.2) |
|  | Stroke | 5 (10.2) | 4 (8.5) | 9 (9.4) |
|  | Chronic pain | 2 (4.1) | 3 (6.4) | 5 (5.2) |
|  | Osteoporosis | 3 (6.1) | 1 (2.1) | 4 (4.2) |
|  | Neurological condition | 1 (2.0) | 0 (0.0) | 1 (1.0) |
|  | Cancer | 0 (0.0) | 1 (2.1) | 1 (1.0) |
|  | Other | 27 (55.1) | 23 (48.9) | 50 (52.1) |
| Smoking status, n (%) | I have never smoked | 16 (32.7) | 22 (46.8) | 38 (39.6) |
|  | I currently smoke | 5 (10.2) | 7 (14.9) | 12 (12.5) |
|  | I am an ex-smoker | 28 (57.1) | 18 (38.3) | 46 (47.9) |
| Alcohol intake<br>(3+ units daily), n (%) | Yes | 7 (14.3) | 6 (12.8) | 13 (13.5) |
|  | No | 42 (85.7) | 41 (87.2) | 83 (86.5) |
| Post-16 education, n (%) | Yes | 29 (59.2) | 32 (68.1) | 61 (63.5) |
| Degree or equivalent, n (%) | Yes | 18 (36.7) | 19 (40.4) | 37 (38.5) |
| Marital status, n (%) | Single | 1 (2.0) | 0 (0.0) | 1 (1.0) |
|  | Divorced/separated | 11 (22.4) | 9 (19.1) | 20 (20.8) |
|  | Widowed | 11 (22.4) | 10 (21.3) | 21 (21.9) |
|  | Cohabiting | 0 (0.0) | 1 (2.1) | 1 (1.0) |
|  | Civil Partnership | 0 (0.0) | 0 (0.0) | 0 (0.0) |
|  | Married | 26 (53.1) | 27 (57.4) | 53 (55.2) |
| Number of children, n (%) | 0 | 3 (6.1) | 3 (6.4) | 6 (6.3) |
|  | 1 | 7 (14.3) | 8 (17.0) | 15 (15.6) |
|  | 2 | 24 (49.0) | 16 (34.0) | 40 (41.7) |
|  | 3 | 10 (20.4) | 15 (31.9) | 25 (26.0) |
|  | 4+ | 5 (10.2) | 5 (10.6) | 10 (10.4) |
| **How many people do you share your home with?, n (%)** | Live alone | 22 (44.9) | 18 (38.3) | 40 (41.7) |
|  | 1 person | 26 (53.1) | 25 (53.2) | 51 (53.1) |
|  | 2 people | 1 (2.0) | 3 (6.4) | 4 (4.2) |
|  | 3 people | 0 (0.0) | 1 (2.1) | 1 (1.0) |
|  | 4 or more people | 0 (0.0) | 0 (0.0) | 0 (0.0) |
| **Current circumstance, n (%)** | Social/physical distancing | 29 (59.2) | 24 (51.1) | 53 (55.2) |
|  | Self-isolating without COVID-19 symptoms | 10 (20.4) | 4 (8.5) | 14 (14.6) |
|  | Self-isolating with COVID-19 symptoms | 0 (0.0) | 0 (0.0) | 0 (0.0) |
|  | Shielding | 10 (20.4) | 19 (40.4) | 29 (30.2) |
|  | Other | 0 (0.0) | 0 (0.0) | 0 (0.0) |
| **Adherence to UK Government's guidance in relation to COVID-19 restrictions, n (%)** | All of the time | 31 (63.3) | 32 (68.1) | 63 (65.6) |
|  | Most of the time | 15 (30.6) | 15 (31.9) | 30 (31.3) |
|  | Some of the time | 3 (6.1) | 0 (0.0) | 3 (3.1) |
|  | A little of the time | 0 (0.0) | 0 (0.0) | 0 (0.0) |
|  | None of the time | 0 (0.0) | 0 (0.0) | 0 (0.0) |

LTC, long term condition
<sup>‡</sup>conditions are not mutually exclusive so percentages not expected to sum to 100.

**Table 2:** Patient Reported Outcome Measures

| Outcome Measure | Control | Intervention |
| --- | --- | --- |
| **PHQ-9, n, mean (SD)** |  |  |
| Baseline | 49, 7.5 (6.2) | 47, 6 (5.6) |
| 1-month | 45, 6.3 (5.9) | 45, 4.9 (4.6) |
| 3-month | 44, 5.7 (5.3) | 42, 5.3 (5.4) |
| **PHQ-9 Categories Baseline, n (%)** |  |  |
| Minimal Depression (0-4) | 22 (44.9) | 21 (44.7) |
| Mild Depression (5-9) | 10 (20.4) | 14 (29.8) |
| Moderate Depression (10-14) | 11 (22.4) | 9 (19.1) |
| Moderately Severe Depression (15-19) | 3 (6.1) | 1 (2.1) |
| Severe Depression (20-27) | 3 (6.1) | 2 (4.3) |
| **PHQ-9 Categories 1-Month, n (%)** |  |  |
| Minimal Depression (0-4) | 22 (48.9) | 24 (53.3) |
| Mild Depression (5-9) | 12 (26.7) | 12 (26.7) |
| Moderate Depression (10-14) | 6 (13.3) | 8 (17.8) |
| Moderately Severe Depression (15-19) | 3 (6.7) | 1 (2.2) |
| Severe Depression (20-27) | 2 (4.4) | 0 (0.0) |
| **PHQ-9 Categories 3-Month, n (%)** |  |  |
| Minimal Depression (0-4) | 21 (47.7) | 23 (54.8) |
| Mild Depression (5-9) | 15 (34.1) | 11 (26.2) |
| Moderate Depression (10-14) | 4 (9.1) | 4 (9.5) |
| Moderately Severe Depression (15-19) | 3 (6.8) | 4 (9.5) |
| Severe Depression (20-27) | 1 (2.3) | 0 (0.0) |
| **GAD-7, n, mean (SD)** |  |  |
| Baseline | 49, 5.2 (5.8) | 47, 3.8 (4.8) |
| 1-month | 45, 4.2 (5.1) | 45, 3.6 (4.2) |
| 3-month | 44, 3.7 (5.0) | 42, 3.5 (3.9) |
| **GAD-7 Categories Baseline, n (%)** |  |  |
| Anxiety (0-4) | 31 (63.3) | 35 (74.5) |
| Mild Anxiety (5-9) | 9 (18.4) | 7 (14.9) |
| Moderate Anxiety (10-14) | 4 (8.2) | 3 (6.4) |
| Severe Anxiety (15-21) | 5 (10.2) | 2 (4.3) |
| **GAD-7 Categories 1-Month, n (%)** |  |  |
| Anxiety (0-4) | 31 (68.9) | 30 (66.7) |
| Mild Anxiety (5-9) | 8 (17.8) | 10 (22.2) |
| Moderate Anxiety (10-14) | 3 (6.7) | 4 (8.9) |
| Severe Anxiety (15-21) | 3 (6.7) | 1 (2.2) |
| **GAD-7 Categories 3-Month, n (%)** |  |  |
| Anxiety (0-4) | 31 (70.5) | 28 (66.7) |
| Mild Anxiety (5-9) | 9 (20.5) | 9 (21.4) |
| Moderate Anxiety (10-14) | 1 (2.3) | 4 (9.5) |
| Severe Anxiety (15-21) | 3 (6.8) | 1 (2.4) |
| **De Jong Gierveld Loneliness Scale, n, mean (SD)** |  |  |
| Baseline | 49, 5.1 (3.2) | 47, 4.6 (3.5) |
| 1-month | 45, 4.6 (3.1) | 45, 4.7 (3.0) |
| 3-month | 44, 5.0 (3.0) | 42, 4.1 (2.9) |
| **De Jong Gierveld Emotional Loneliness Subscale, n, mean (SD)** |  |  |
| Baseline | 49, 3.1 (1.9) | 47, 3.0 (2.0) |
| 1-month | 45, 3.0 (1.7) | 45, 3.1 (1.9) |
| 3-month | 44, 3.4 (1.7) | 42, 3.0 (1.7) |
| **De Jong Gierveld Social Loneliness Subscale, n, mean** |  |  |
| (SD) |  |  |
| Baseline | 49, 2.0 (1.8) | 47, 1.6 (1.8) |
| 1-month | 45, 1.5 (1.8) | 45, 1.6 (1.8) |
| 3-month | 44, 1.6 (1.8) | 42, 1.1 (1.6) |
| **SF-12v2 (Physical Component Score), n, mean (SD)** |  |  |
| Baseline | 49, 39.4 (10.7) | 47, 40.3 (11.3) |
| 1-month | 45, 40.0 (10.5) | 45, 41.4 (12.4) |
| 3-month | 44, 41.0 (11.5) | 42, 41.8 (11.7) |
| **SF-12v2 (Mental Component Score), n, mean (SD)** |  |  |
| Baseline | 49, 47.0 (13.9) | 47, 48.9 (10.5) |
| 1-month | 45, 48.4 (13.0) | 45, 52.0 (9.5) |
| 3-month | 44, 49.0 (11.5) | 42, 51.1 (9.7) |

### Engagement with the BASIL intervention

Levels of engagement with the Behavioural Activation intervention were high. Of the 47 intervention participants randomised to the Behavioural Activation intervention group, 45 (95.7%) commenced the intervention, with 44 participants completing two or more sessions. The number of sessions completed range from 0 to 8 (median of 6 sessions) out of a total of up to 8 sessions. Participants preferred telephone over video contact. Sessions lasted an average of 36.7 minutes (SD 15.7). Two participants withdrew from the intervention (after completing one and two sessions, respectively); one participant stated their reason for withdrawal was that they felt ‘well’ and the intervention was ‘not for them’ as they were already engaging in BA-related activities. At one month (the primary clinical outcome point), the median number of completed sessions for people receiving the Behavioural Activation intervention was 3, and almost all participants were still receiving the BA intervention.

### Retention, follow up, withdrawal and completeness of data

Of the 96 participants randomised into the study, 90 (93.8%) completed the one month follow-up, and 86 (89.6%) completed the three month follow-up. Reasons for withdrawal include personal reasons, family bereavement, and finding the study/study questions upsetting and anxiety-provoking.

Data completeness was good with all patient-reported outcome measures (PHQ-9, GAD-7, De Jong Gierveld Loneliness Scale and SF-12v2).

### Outcome data and between group comparisons at 1 and 3 months

Unadjusted between-group mean differences tended to favour the intervention across measures and timepoints. The adjusted mean difference (AMD) between groups in the PHQ-9 favoured the intervention group at one month (−0.50, 95% CI −2.01 to 1.01), and the usual care group at three months (0.19, 95% CI −1.36 to 1.75) (Table 3). In De Jong Gierveld score, the adjusted mean difference favoured the usual care group at one month (0.28, 95% CI −0.51 to 1.06), and there was a statistically significant benefit for the intervention group at three months (−0.87, 95% CI −1.56 to −0.18) (Table 3).

**Table 3.** Adjusted mean difference between groups in PHQ-9 and De Jong Gierveld scale at one and three months post-randomisation

| Outcome measure | Adjusted mean difference (95% CI) |
| --- | --- |
| *1-month* |  |
| PHQ-9 | -0.50 (-2.01 to 1.01) |
| De Jong Gierveld scale | 0.28 (-0.51 to 1.06) |
| *3-month* |  |
| PHQ-9 | 0.19 (-1.36 to 1.75) |
| De Jong Gierveld scale | -0.87 (-1.56 to -0.18) |

### Process evaluation and changes to the intervention in light of participant feedback

Intervention participants were invited to be interviewed following completion of the one month follow up and conclusion of their participation in the BASIL Pilot intervention. Semi-structured interviews were conducted with 15 participants who completed the BA intervention. A study participant who did not complete the BASIL intervention was interviewed as a ‘non-completer’. All nine BSWs who delivered the BASIL intervention were interviewed between August 2020 and November 2020. All interviews were conducted over the telephone, digitally-recorded with consent and transcribed verbatim. Transcripts formed the basis for analysis. We summarise the key findings from a thematic analysis34 and subsequent changes to the BASIL intervention ahead of the BASIL Main Trial. Analysis sensitised by the ‘Theoretical Framework of Acceptability (TFA;35) will be reported separately. Intervention participant and BSW demographics are reported in the appendices.

### Summary of findings

#### Recruitment and study eligibility

Study recruitment methods appeared to be generally acceptable and clear:

> *‘the doctor gives you a warning that this is about to happen [be contacted] and then you’re prepared when somebody phones up that it’s not a scam, a con, which is what I don’t worry about it because I know how to deal with it but to some people it could be worrying’ (OA16)*

Some participants, generally participants without symptoms of depression at study entry and also some BSWs, raised the importance of more targeted recruitment to the BASIL intervention:

> *‘I do think just some consideration needs to be given to who we’re targeting, maybe it’s not quite so useful for people on the threshold of depression and feel that they’re doing quite well (BSW 04)*
>
> *I think possibly it needs to be more targeted, so anybody who has a painful medical condition or who lives alone who is isolated, certainly I think it would benefit them a lot. I think the wide spread that you’ve currently can be more targeted and more focused and more helpful to more people in that sense (OA02)*

#### Intervention delivery and content

Remote delivery of the intervention by telephone was acceptable. Although video calls were offered, these were not taken up by participants. Some participants and BSWs reported they would have preferred face-to-face intervention delivery, had this been possible. One participant suggested that those with hearing difficulties would find telephone delivery more difficult. The number and frequency of intervention sessions was acceptable, although one participant reported that they would prefer one session per week, to allow them time to implement agreed activities and plans. Some BSWs reported it could be difficult to stick to the 30-minute timing for more complex or isolated cases, where meetings took longer.

The BASIL Behavioural Activation self-help booklet was thought to be engaging, and people found the mood/behaviour cycle understandable. However, some participants-those with few depression symptoms at study entry-found this model of limited relevance. Several participants reported that they would use the booklet after the intervention ended:

> *‘So, in days of darkness I’ll be able to flick through it [the booklet] and say, that’s what that was all about, how to break things down and not get upset about them and not let them get you down’ (OA06)*

The patient stories in the booklet were reported to be relevant, although some participants reported that booklet activity examples could ideally be more varied. Both BSWs and participants found activity planning to be sometimes difficult, especially where some services were shut under lockdown conditions. Planned activities may therefore need to be sufficiently flexible to accommodate changes in COVID-19 restrictions.

#### Study adaptations

The process evaluation led to intervention adaptations for the BASIL main trial, including: refining the study eligibility criteria, adaptions to the self-help booklet to make reference to a wider range of example activities, making reference to modifying goals, bringing discussion of ‘functional activities’ forward and providing a large print version of the self-help booklet when needed.

## Discussion

### Main findings

The BASIL trial is an external pilot trial, designed to test acceptability of an adapted intervention and to refine trial procedures and design prior to undertaking a full scale trial.21,36 Our main finding is that higher risk older people with long-term conditions living under COVID restrictions were receptive to an approach to participate in a trial of a behavioural intervention. When offered BA they preferred telephone contact rather than an offer of technology-enabled video calling. Levels of engagement with BA were high, with a greater proportion completing six or more planned sessions. Some people with long-term conditions declined the BASIL offer of telephone support. In qualitative interviews it was clear that those with very low levels of depression and good adaptation to socially-isolating restrictions were not an appropriate target group. This has led us to refine and target our intervention in a fully powered trial, and we will now only focus on older people who have some depressive symptoms above a threshold, and at risk of further deterioration in mental health.

We also sought to establish the ideal point at which to judge the short-run outcome of the intervention under a fair test. At the candidate primary outcome (one month) only a small number of BA sessions had been completed; whereas at three months the intervention had been completed by all engaged participants. This has led us to use 3 months as the follow-up for the primary outcome in the main trial.

Although underpowered to test effectiveness, the between-group comparisons using confidence intervals included benefit for BA in mitigating levels of depression at 3 months. For our measure of loneliness, there was a significant benefit which excluded the null and was unlikely to be a chance finding. Our preliminary analysis is in line with a confidence interval approach to the interpretation of pilot trials37 and we are keen not to overinterpret the positive finding of mitigating loneliness using BA. However, this is an encouraging finding which justifies the need for a full scale trial, where the consistency of this effect will be tested with greater levels of power and precision. The BASIL+ trial (the fully powered follow-on trial) is now underway and is pre-registered (https://doi.org/10.1186/ISRCTN63034289) to reflect the design adaptations from the pilot study.

### Strengths and limitations of the BASIL pilot study and comparison with other studies

The BASIL trial and nested qualitative work adds to an emerging literature on the use of psychological interventions that incorporate cognitive or behavioural strategies to address loneliness and its causal role in depression.38 Research to date has shown behavioural approaches to be highly effective in the treatment of depression among older people17,19,39 and the preliminary results of the BASIL trial lend support to this approach in the face of COVID restrictions. A fully-powered trial of BA is now underway, and in time this will report on the short-and long-term clinical and cost effectiveness of a scalable behavioural psychosocial intervention. This will add to an emerging trial based literature to establish ‘what works’ in the mitigation of loneliness.11,40

Our pilot trial was also undertaken very rapidly and in response to the fast-moving COVID pandemic in early 2020. As such we, along with other researchers undertaking trials during COVID, have had to adapt the methods used to generate randomised evidence. We have shown that it is possible to deliver trials with adaptations to minimise patient contact and streamline recruitment procedures. This makes us confident that this is an efficient method of participant engagement and follow-up for future trials, both under COVID and beyond the pandemic. It is of note that the time elapsed between the onset of the pandemic and the recruitment of the first participant was less than 3 months. Finally, we have chosen to study the impact of a plausible psychosocial intervention to mitigate depression and loneliness in an at-risk population of older people with multimorbidity. Population surveys under COVID-19 have shown that younger people are also at risk of loneliness41 and psychological deterioration.42 It is important that interventions to tackle the higher rates of depression and loneliness in all age groups are also developed and evaluated.

### Conclusions and modifications to the design of the BASIL full trial

At the outset of the COVID-19 it was predicted that there would be significant impacts on public mental health,5 including loneliness and depression as a consequence of pandemic restrictions. This has come to pass42 and population surveys indicate increased reports of loneliness and reports of depression.2 The pandemic has also prompted a number of studies to understand the impacts of COVID-19,43 but there have been very few studies to evaluate psychosocial interventions to mitigate psychological impact. To our knowledge, BASIL is the first study to report trial-based evidence.

The preliminary results are in line with potential benefit for this intervention at 3 months, and we will now test the short-and long-term clinical and cost effectiveness. This evidence may prove to be useful in improving the mental health of populations during the time of COVID-19 and also in mitigating depression and loneliness in socially isolated at-risk populations after the pandemic has passed.14

## Data Availability

Anonymised data will be made available upon reasonable request, which must include a protocol and statistical analysis plan and not be in conflict with our prespecified publication plan, consistent with our data sharing policy (available on request from SG). The BASIL research collective is especially keen that the BASIL data contributes to prospective meta-analyses and individual patient data meta-analyses. Requests for data sharing will be considered by SG and the independent trial steering and data monitoring committee.

## Contributions of the authors

SG, DE, CCG, EL, DMcM, CH, DB and SGa planned the trial, contributed to the trial design and drafted the trial protocol. SG and DE led manuscript writing, managed the trial as chief investigators, and critically revised the manuscript. SG, EL, DMcM, CCG, CH, PC, GTT, AC, TG, AHi, KL, SDS, TO and JW contributed to trial design and trial management meetings.

SG, CCG, DE, DMcM and DB designed the intervention and BSW training materials, and DB, DMcM, CCG and DE delivered the BSW training. EL led the day-to-day management of the trial, and SGa and RW were the trial coordinators. DB, SC and DMcM provided BSW clinical supervision. SGa, LB, AH, ER, LS and RW facilitated participant recruitment and follow-up data collection, and participated in trial management meetings. ER and LS delivered the BA intervention. CF, KJ and CH developed the statistical analysis plan and analysed the quantitative data. CS, CCG and PC led the process evaluation. CS conducted interviews, and CS, CCG, PC and AH analysed the qualitative data. All authors contributed to the drafts of manuscripts and read the final manuscript. The York Trials Unit act as data custodians.

## Acknowledgements

We would like to thank: the participants for taking part in the trial, general practice and North East and North Cumbria Local Clinical Research Network staff for identifying and facilitating recruitment of participants, the independent Programme Steering Committee members for overseeing the study, and our PPI AG members for their insightful contributions and collaboration.

## Appendix

**Older adult Demographics (Completer participants)**

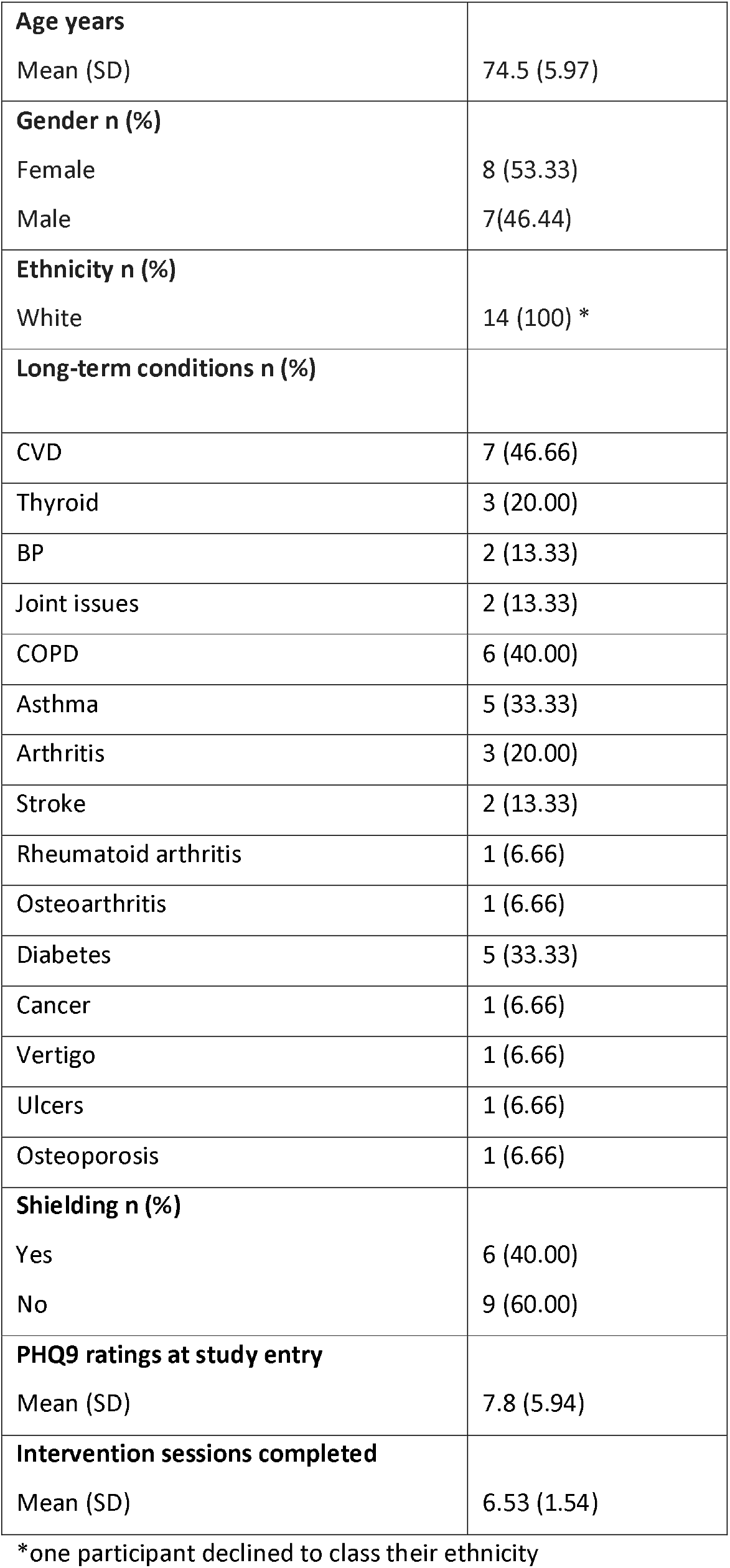

**Older adult Demographics (non-completer participants)**

|  |  |
| --- | --- |
| **Age years** | 80 |
| **Gender** |  |
| Female | 1 |
| **Ethnicity n** |  |
| White | 1 |
| **Long-term conditions n (%)** |  |
| CVD | 1 (100) |
| Chronic pain | 1 (100) |
| **Shielding n** |  |
| No | 1 |
| **PHQ9 ratings at study entry** |  |
| Number | 1 |
| **Intervention sessions completed** |  |
| Number | 1 |

**Table:** BASIL Support Worker Demographics

Table: **BASIL Support Worker Demographics**
|  |  |
| --- | --- |
| **Age years** |  |
| Mean (SD) | 40.16, (14.01) * |
| **Gender n (%)** |  |
| Female | 7 (100) * |
| **Ethnicity n (%)** |  |
| White | 7 (100) * |
| **Job role n (%)** |  |
| Retired community mental health nurse | 1 (11.11) |
| Research and registered mental health nurse | 1 (11.11) |
| Psychological wellbeing practitioner | 3 (33.33) |
| Assistant psychologist | 1 (11.11) |
| Research Assistant | 3 (33.33) |
| Retired community mental health nurse | 1 (11.11) |
| **Years of experience in current and related roles** |  |
| Mean (SD) | 16.61 (13.39) |

\*Although nine BSWs took part in interviews, only 7 BSWs provided demographics for gender and ethnicity and 6 BSWs provided demographics for age. These numbers represent the age, gender and ethnicity categories above.

